# Retrospective Analysis of Neonatal Hyperbilirubinemia at Hiwot Fana Specialized Hospital, Ethiopia: Prevalence, Associated Factors, and Treatment Outcomes

**DOI:** 10.1101/2024.05.13.24307293

**Authors:** Mohamed Abdirahman Shugri, Temesgen Teferi Libe, Feven Mekonnen Gebru

**Affiliations:** School of Medicine, College of Health and Medical Sciences, Jigjiga University, Eastern, Ethiopia; Department of Pediatrics and Child Health, College of Health and Medical Sciences, Haramaya University, Harar, Ethiopia; Abinet Health Centre, Addis Abeba Health bureau, Addis Abeba, Ethiopia

**Keywords:** Neonatal hyperbilirubinemia, Prevalence, Associated factors, Treatment outcomes, Eastern Ethiopia

## Abstract

**Background:** Neonatal hyperbilirubinemia, also known as neonatal jaundice, is a condition characterized by yellowish discoloration of the skin and eyes caused by bilirubin buildup in the body. Understanding its prevalence, associated factors, and outcomes is crucial for effective management.

**Objectives:** This study aimed to assess the prevalence, associated factors, and treatment outcomes of neonatal hyperbilirubinemia at Hiwot Fana Specialized Hospital, Eastern Ethiopia.

**Methodology:** A hospital-based cross-sectional study was conducted from December 1, 2020, to December 30, 2020, involving 328 neonates admitted to the Neonatal Intensive Care Unit. Data were collected using a checklist and analyzed using SPSS version 22.

**Result:** Neonatal hyperbilirubinemia prevalence was 44.2%, with 1.4% progressing to bilirubin encephalopathy. Sepsis (44.8%) and prematurity (22%) were major contributing factors. Most neonates were treated with phototherapy alone (82.3%), and 93.8% showed improvement.

**Conclusion:** The study underscores the importance of early detection and management of neonatal hyperbilirubinemia to prevent adverse outcomes. Recommendations include integrating jaundice prevention into antenatal care and ensuring timely bilirubin level checks in neonates.

## 1. INTRODUCTION

### 1.1 Background

Neonatal hyperbilirubinemia, also known as neonatal jaundice, is a condition characterized by a yellowish discoloration of the skin, sclera, and mucous membranes, stems from the accumulation of bilirubin in the body [1]. This condition manifests when serum bilirubin levels exceed 85μmol/l (5mg/dl), primarily due to heightened bilirubin production from senescent fetal red blood cells and the infant’s limited capacity to eliminate bilirubin effectively [2].

Of significant concern is the potential for kernicterus, a form of bilirubin encephalopathy, which arises from unconjugated hyperbilirubinemia. Kernicterus can result from hemolytic diseases like Rh incompatibility or deficiencies in the liver’s ability to conjugate bilirubin due to enzymatic defects[3]. Its clinical manifestations span from acute to chronic phases, with symptoms such as hypotonia, poor feeding, high-pitched cry, and, in severe cases, cerebral palsy, sensory-neural deafness, seizures, and neurocognitive impairment [4].

Globally, neonatal jaundice affects a substantial proportion of newborns, with up to 60% of term and 80% of preterm infants experiencing it within their first week of life [5].

Studies across various regions have reported varying incidences of neonatal hyperbilirubinemia, with Pakistan and Nigeria documenting rates of 27.6% and 19.6%, respectively [6].

Contributing to these elevated bilirubin levels are factors such as infection, ABO incompatibility, and glucose-6-phosphate dehydrogenase (G6PD) deficiency [7].

In Ethiopia, neonatal jaundice poses a significant threat, with prevalent factors including neonatal sepsis, preterm birth, hypoxia, and acidosis exacerbating the risk of acute bilirubin encephalopathy [8]. Despite its prevalence and associated morbidity, there’s a scarcity of comprehensive studies exploring the burden and determinants of neonatal jaundice in Ethiopia, particularly in the study area of Hiwot Fana Specialized University Hospital, Eastern Ethiopia.

This study seeks to address this gap by investigating the incidence of neonatal jaundice and its associated factors among neonates admitted to the neonatology unit at Hiwot Fana Specialized University Hospital. Understanding the epidemiology and determinants of neonatal jaundice in this context is crucial for several reasons.

Firstly, it provides vital insights for hospital protocols, enabling the development of targeted strategies for the prevention and management of neonatal hyperbilirubinemia. By identifying modifiable risk factors and early treatment options, healthcare providers can optimize care delivery and improve neonatal outcomes within the hospital setting.

Secondly, the findings of this study have broader implications for regional health policies and programs. Harari Regional Health Bureau and other stakeholders can leverage this data to formulate evidence-based interventions aimed at reducing the burden of neonatal jaundice and improving neonatal health outcomes in the region.

Lastly, this study contributes to the existing body of knowledge on neonatal jaundice, serving as a resource for future research endeavors in this field. Researchers and practitioners alike can build upon these findings to deepen our understanding of the epidemiology, etiology, and management of neonatal jaundice, both within Ethiopia and beyond.

### 1.2 Objectives

#### General objective

To assess prevalence, associated factors, and treatment outcome of Neonatal Hyperbilirubinemia among neonates admitted to Hiwot Fana Specialized University Hospital, Eastern Ethiopia from January 2019 to December 2020.

#### Specific Objectives

1. To determine prevalence of Neonatal Hyperbilirubinemia among neonates admitted to Hiwot Fana Specialized Hospital.
2. To assess factors associated with Neonatal Hyperbilirubinemia among neonates admitted to Hiwot Fana Specialized Hospital
3. To determine treatment outcome of Neonatal Hyperbilirubinemia among neonates admitted to Hiwot Fana Specialized Hospital.

## 2. Methodology

### 2.1 Study area and period

This study took place at Hiwot Fana Comprehensive Specialized Hospital (HFCSH), a teaching hospital affiliated with Haramaya University in Harar town, Harari Region, Ethiopia. HFCSH serves as a referral center for Eastern Ethiopia, catering to a population of approximately 5.8 million people, including residents of the Harari Region, Dire Dawa City Administration, Somali Region, and the Eastern Hararghe Zone of the Oromia Regional State. According to unpublished Health Management Information System data from 2020, HFCSH admits around 3,000 pediatric patients annually, half of whom are neonates. Additionally, the hospital conducts approximately 5,500 deliveries each year.

The study was conducted from December 1, 2020, to December 30, 2020.2.2.

### 2.2. Study design

A hospital-based cross-sectional study design was employed.

### 2.3 populations

#### 2.3.1 Source population

The source population included all neonates admitted to the Neonatal Intensive Care Unit (NICU) of HFCSH from January 1, 2019, to December 30, 2020.

#### 2.3.2 Study population

The study population comprised selected neonates among those admitted to the NICU of HFCSH during the same period.

### 2.4. Inclusion and exclusion criteria

#### 2.4.1 Inclusion Criteria

All neonates of age group from birth to 28 days who were admitted to NICU of Hiwot Fana Specialized Hospital during the study period.

#### 2.4.2 Exclusion Criteria

Patients diagnosed with neonatal jaundice but with incomplete charts or without a recorded serum bilirubin level were excluded from the study.

### Sample size determination and sampling technique

#### 2.5.1 Sample Size determination

##### Objective 1

Prevalence of Neonatal Jaundice

A single population proportion formula was employed, considering a prevalence (p) of 44.9% from a previous study conducted at Black Lion Hospital (Kassa et al., 2018). We aimed for a 95% confidence interval and a 5% degree of precision. To account for a potential 5% non-response rate, the initial sample size was adjusted accordingly. This calculation resulted in a final sample size of 328.

##### Objective 2

Associated Factors of Neonatal Jaundice

For the second objective, which focused on identifying factors associated with neonatal jaundice, a double population proportion formula was used. Sample sizes for specific factors were calculated using OpenEpi data statistical software version 3.1. The calculations assumed a 95% confidence level, 80% power, and a ratio of unexposed to exposed neonates close to 1 (data from different literature sources was used for these calculations). (Table 1)

**Table 1:** Sample size determinations using double population proportions formula for the second objective.

| Factors | Neonatal jaundice |  | Odds ratio | Non-response rate | Sample size | Reference |
| --- | --- | --- | --- | --- | --- | --- |
|  | Exposed | Non-exposed |  |  |  |  |
| Sepsis | 49 | 27 | 2.64 | 10% | 170 | [9] |
| Blood group incompatibility | 84.8 | 23.9 | 18.21 | 10% | 26 | [9] |
| Duration of labor | 34 | 44 | 4.39 | 10% | 76 | [10]. |
| Place of delivery | 36.4 | 44.3 | 4.37 | 10% | 78 | [10]. |

The larger sample size, 328, was taken as final sample size.

#### 2.5.2 Sampling techniques

A systematic sampling method was employed, selecting every 6th value among neonates admitted to the NICU of HFCSH from January 1, 2019, to December 30, 2020.

### 2.6. Data collection method

#### 2.6.1. Data collection tool and procedure

A checklist developed in the English language, containing variables to be measured, was utilized. The checklist was adapted from various literature sources [9, 10]. Neonatal admission cards from the NICU of HFCSH were isolated and counted, and their numbers were arranged in the order of admission. Data collection involved systematically collecting necessary information from every 6th value in the order.

#### 2.6.2 Data collectors

Two BSc nurses and one resident were trained and recruited as data collectors under the supervision of the principal investigator.

### 2.7 Study variable

#### 2.7.1 Dependent variables

- Neonatal jaundice.
- Treatment outcome

#### 2.7.2 Independent variables

- Maternal factors: blood group and Rh, antenatal care (ANC) follow-up.
- Neonatal factors: age, gestational age, birth weight, blood group and Rh, hematocrit, total serum bilirubin (TSB), mode of delivery, birth trauma, breast milk jaundice, sepsis, breastfeeding.

### 2.9. Data quality control

Measures were taken to ensure the validity and reliability of data collection, including training of data collectors, pre-testing of the questionnaire, supervision during data collection, and double data entry with consistency checks.

### 2.10. Methods of data analysis

The collected data underwent a thorough cleaning process to ensure completeness before being entered into EPI Data version 8.1. This process involved coding, cleaning, and editing to minimize logical errors and prevent skipping patterns in the data entry. Subsequently, the data was exported to SPSS version 22 for analysis. Descriptive statistics were employed to summarize the data. This included calculating proportions and other summary measures, which were then presented using simple frequencies, tables, and figures.

To investigate the associations between independent and dependent variables, both bivariate and multivariate analyses were conducted using binary logistic regression. The assumptions for the binary logistic regression model were verified. Goodness-of-fit was assessed using the Hosmer-Lemeshow statistic and omnibus tests.

Variables with a p-value less than 0.25 in the bivariate analysis were included in the final model of the multivariate analysis. This approach helped control for potential confounding factors. The direction and strength of the statistical associations were measured by calculating odds ratios with 95% confidence intervals (CI). Multivariate analysis using binary logistic regression was employed to estimate adjusted odds ratios (also with 95% CI) to identify independent predictors for neonatal jaundice and its associated factors. In this study, a p-value less than 0.05 was considered statistically significant.

### 2.11. Ethical consideration

Ethical approval was obtained from the Institutional Research Ethics Review Committee (IHRERC) of Haramaya University. Informed consent was obtained from participants, and confidentiality was maintained throughout the study.

## 3. RESULT OF THE STUDY

### 3.1. Socio-demographic characteristics of neonates in the study, HFSUH

In a retrospective cross-sectional study, 328 neonates were systematically selected from all neonates admitted to the NICU of HFSUH from January 1, 2020, to December 30, 2020, to evaluate the magnitude and associated factors of neonatal hyperbilirubinemia. Of these, 176 (53.7%) were male and 152 (46.3%) were female. Among the neonates included, 145 (44.2%) developed neonatal hyperbilirubinemia. At the time of admission, 138 (42%) neonates were aged 3-6 days, and 247 (75.3%) weighed between 2500gm-4000g. The duration of hospital stays ranged from 1 to 31 days, with 211 (64.3%) discharged within a week and 10 (10.6%) discharged after 21 days. Premature neonates accounted for 116 (35.4%), and 60 (18.3%) neonates’ mothers did not know their gestational age. (Table 2)

**Table 2:** Socio-demographic characteristics of Admitted Neonates.

| Socio-demographic characteristics | Category | Frequency/percent; among all neonates (328) | Frequency(percent); Jaundiced Neonates |
| --- | --- | --- | --- |
| Sex | Male | 176(53.7%) | 89(61.4%) |
|  | Female | 152(46.3%) | 56(38.6%) |
| Age at admission (days) | <= 2 | 93(28.4%) | 21(14.5%) |
|  | 3-6 | 138(42%) | 65(44.8%) |
|  | > 6 | 97(29.6%) | 59(40.7%) |
| Weight (grams) | < 2500 | 71(21.6%) | 21(14.5%) |
|  | 2500-4000 | 247(75.3%) | 118(81.4%) |
|  | >= 4000 | 10(3.1%) | 6(4.1%) |
| Gestational age (weeks) | < 37 weeks | 116(35.4%) | 36(24.8%) |
|  | >= 37 weeks | 152(46.3%) | 70(48.3%) |
|  | Unknown | 60(18.3%) | 39(29.9%) |

#### 3.2 Magnitude of neonatal hyperbilirubinemia

Neonatal hyperbilirubinemia was diagnosed in 145 (44.2%) of the reviewed neonates. Only 2 (1.4%) cases progressed to bilirubin encephalopathy. Males had a higher prevalence of hyperbilirubinemia compared to females (61.4% vs. 38.6%). The most common age group for presentation with jaundice was 3-6 days old (44.8%). Neonates weighing 2500-4000gm at admission were most likely to develop jaundice (81.4%). Prematurity was a contributing factor in 35.4% of jaundiced neonates. The majority (60%) of these neonates were discharged within the first week after admission. The most frequent onset of jaundice occurred between 3 and 6 days after birth (44.8%). Less frequent jaundice onset occurred ≤ 3 days (14.5%) or > 6 days (40.7%) after birth.

In terms of serum bilirubin levels, 33 (22.8%) neonates had levels ≥20mg/dL, while 83 (57.2%) had levels between 10mg/dL-20mg/dL. Notably, both neonates with bilirubin encephalopathy had serum bilirubin levels of 28mg/dL and 25mg/dL. (Table 2)

### 3.3. Associated factors of neonatal hyperbilirubinemia

Sepsis (44.8%) and prematurity (22.0%) were the major causes of neonatal hyperbilirubinemia in this study. Hemolytic disease caused by Rh incompatibility (4.8%) and ABO incompatibility (10.3%) were also identified as contributing factors (Table 3). Multiple contributing factors were identified in 16 (11.0%) neonates with elevated bilirubin levels. The most common combinations were ABO incompatibility combined with prematurity (6.2%) and sepsis combined with prematurity (3.8%). Both neonates with bilirubin encephalopathy had multiple contributing factors.

**Table 3:**
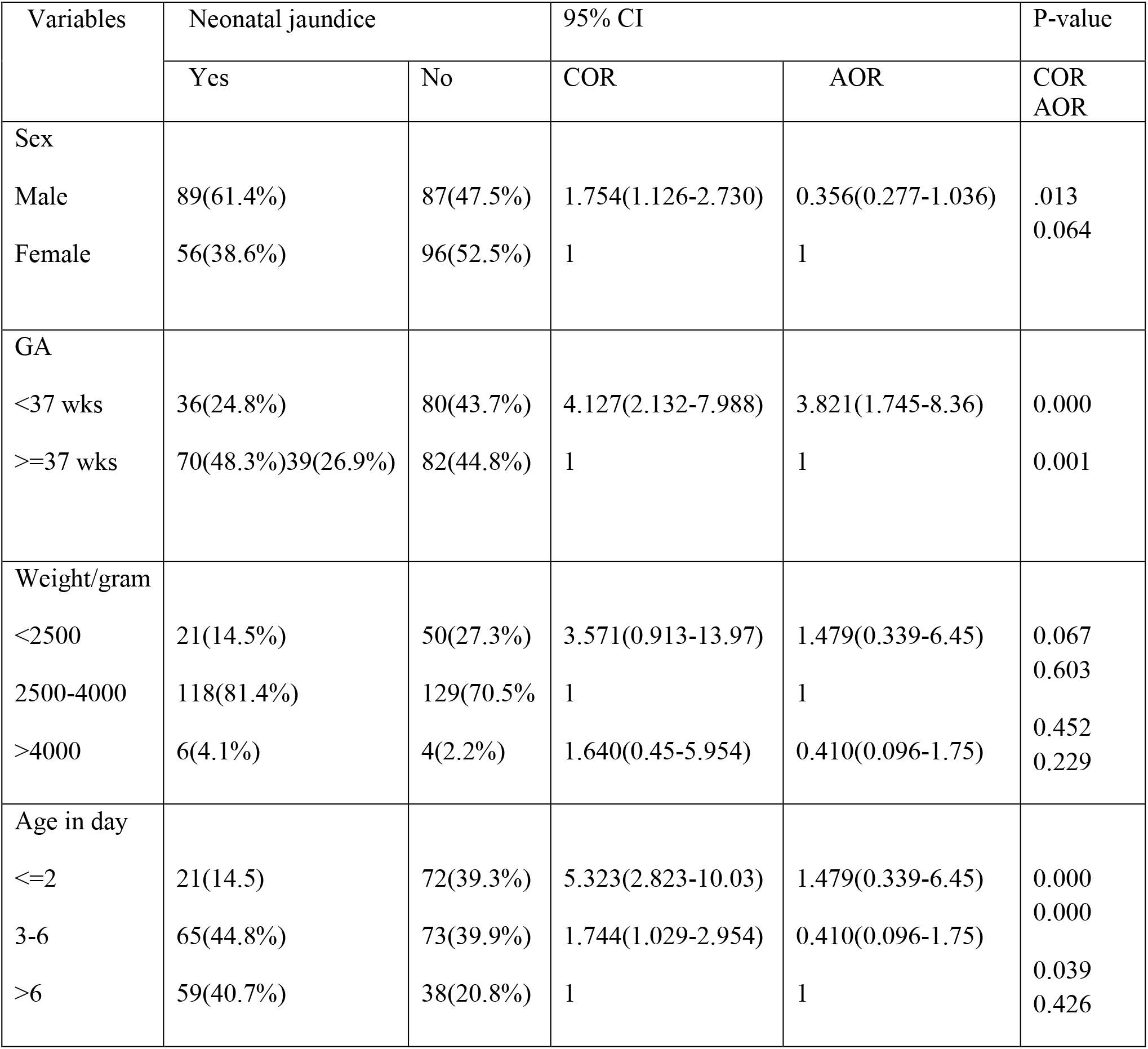
Bivariate and multivariate regression for associated factors with neonatal hyperbilirubinemia.

Sepsis was a leading cause of neonatal hyperbilirubinemia; however, blood cultures were only performed in 2 neonates, and both cultures were positive.

Binary logistic regression analysis identified several factors significantly associated with the development of neonatal hyperbilirubinemia. Neonates admitted at ≤ 2 days old were 5.3 times more likely (OR = 5.323, 95% CI: 2.823-10.039, p-value=0.000) to develop hyperbilirubinemia compared to those admitted after 6 days old. Similarly, neonates with a birth weight less than 2500 grams were 3.6 times more likely (OR = 3.571, 95% CI: 0.913-13.970, p-value=0.067) to experience hyperbilirubinemia compared to those weighing 4000 grams or more. Premature birth (gestational age less than 37 weeks) was also a significant risk factor, with neonates in this group being 4.1 times more likely (OR = 4.127, 95% CI: 2.132-7.988, p-value=0.000) to develop hyperbilirubinemia compared to neonates with unknown gestational age.

However, multivariate logistic regression analysis only confirmed a significant association between neonatal hyperbilirubinemia and gestational age. Premature birth (gestational age less than 37 weeks) remained a significant risk factor, with neonates in this group being 3.8 times more likely (AOR = 3.821, 95% CI: 1.745-8.368, p-value=0.001) to develop hyperbilirubinemia compared to those born at 37 weeks or later (Table 3)

### 3.4. Condition at discharge and management of neonates with hyperbilirubinemia

Most jaundiced neonates were treated with either phototherapy alone (82.3%) or a combination of phototherapy and blood transfusion (7.6%). Two neonates with planned exchange transfusions did not receive them due to a blood shortage at the hospital at the time of admission. One of these neonates with bilirubin encephalopathy died. Neonatal death due to bilirubin encephalopathy occurred in 2 cases (1.4%) among those with hyperbilirubinemia. Overall, 93.8% of jaundiced neonates improved and were discharged home. However, 7 (4.8%) left against medical advice.

## 4. DISCUSSION

This study revealed a high prevalence of neonatal hyperbilirubinemia, with 145 (44.2%) cases identified, of which 2 (1.4%) neonates developed bilirubin encephalopathy. These findings were comparable to other studies conducted in Ethiopia, which reported a prevalence of 37.3% [10] and 44.9% [9]. However, the prevalence in this study was significantly higher compared to studies from other parts of the world, such as 13.3% *[11]*, 27.6% (*[6]* and 24.8% [12]. This disparity may be attributed to differences in study areas or socio-demographic characteristics.

The study also found that neonatal death due to neonatal hyperbilirubinemia was 2 (1.4%), both of which were attributed to bilirubin encephalopathy. This underscores the seriousness of hyperbilirubinemia, leading to long-term sequelae in survivors and death in neonates. These findings align with research by Aboyans and Collaborators [13], which highlighted the increasing recognition of neonatal hyperbilirubinemia as a significant contributor to global neonatal mortality.

Furthermore, significant associations were found between neonatal hyperbilirubinemia and gestational age. The study revealed that 70.4% of neonates developed hyperbilirubinemia in the first week of life. Similar findings were observed in studies conducted in various regions, indicating a consistent trend of neonatal hyperbilirubinemia onset within the early postnatal period. Among the associated factors identified in this study, sepsis (44.8%) and prematurity (22%) were prominent contributors to neonatal hyperbilirubinemia. Other contributing factors included ABO incompatibility (10.3%), Rh incompatibility (4.8%), breastfeeding (8.9%), and other known causes (15.9%).

Comparisons with similar studies revealed similarities and differences in associated factors. For instance, a study conducted at Black Lion Hospital in 2017 found ABO incompatibility (35.6%) and sepsis (18.8%) to be major associated factors. Meanwhile, a study in West India University in 2012 identified ABO incompatibility (35%), prematurity (11%), Rh incompatibility (3.5%), and idiopathic causes (9%) as significant factors. The prevalence of ABO incompatibility was consistent across studies, suggesting its importance in neonatal hyperbilirubinemia.

Additionally, challenges in early detection of hyperbilirubinemia were noted, particularly in neonates whose mothers lacked antenatal follow-up and did not know their gestational age. This could lead to delays in diagnosis and increased complications. Moreover, the study highlighted the association between elevated serum bilirubin levels and the presence of multiple associated factors, indicating a cumulative effect on bilirubin levels

In terms of management, the majority of jaundiced neonates received phototherapy alone, while a smaller proportion required a combination of phototherapy and blood transfusion. Challenges in access to exchange transfusion due to blood shortages were noted, underscoring the need for improved healthcare infrastructure and resources.

### Limitation and the Strength of the study

#### Strength

This study benefited from well-maintained medical records, allowing researchers to obtain accurate information related to neonatal hyperbilirubinemia.

#### Limitation

However, the retrospective nature of the study limited the ability to locate and follow up with discharged subjects to determine if they developed long-term complications. Additionally, the study’s single-site design and limited sample size restrict the generalizability of the findings to other institutions.

## 6. CONCLUSION

The study findings highlight the significant burden of neonatal hyperbilirubinemia at Hiwot Fana Specialized University Hospital, Eastern Ethiopia. Sepsis, prematurity, and other associated factors contribute to the high prevalence of this condition. Effective management strategies, including phototherapy and blood transfusions, play a crucial role in improving outcomes for affected neonates. However, challenges such as blood shortages pose barriers to optimal care delivery.

## Data Availability

Any of the data used for analysis in the study is available from the corresponding author and ready to be provided up on reasonable request.

## AKNOWLEGMENTS

We would like to express our sincere gratitude to Haramaya University for their invaluable support. This includes both the ethical clearance they provided and the technical and financial assistance they offered.

## Contribution of authors

Conceptualization: Mohamed A Shugri, Temesgen T Libe, Feven M Gebru; formal analysis: Mohamed A Shugri, Temesgen T Libe, Feven M Gebru; investigation: Mohamed A Shugri, Temesgen T Libe; methodology: Mohamed A Shugri, Temesgen T Libe, Feven M Gebru; Validation: Mohamed A Shugri, Temesgen T Libe; writing the original draft: Mohamed A Shugri, Feven M Gebru.

